# United States Marijuana Legalization and Opioid Mortality Trends Before and During the First Year of the COVID-19 Pandemic

**DOI:** 10.1101/2023.10.03.23296502

**Authors:** Archie Bleyer, Brian Barnes, Kenneth Finn

## Abstract

To determine if marijuana legalization reduced opioid mortality, the U.S. opioid and fentanyl subset death trends during the 2010-2019 decade were compared in states and District of Columbia (D.C.) (jurisdictions) that had implemented marijuana legalization with states that had not. Acceleration of opioid mortality during 2020, first year of the COVID-19 pandemic, was also compared in recreational and medicinal-only legalizing jurisdictions. Joinpoint methodology was applied to Centers for Disease Control and Prevention WONDER data. Trends in legalizing jurisdictions were cumulative aggregates. The overall opioid and fentanyl death rates and percentage of opioid deaths due to fentanyl increased more during 2010-2019 in jurisdictions that legalized marijuana than in those that did not (pairwise comparison p = 0.007, 0.05, and 0.006, respectively). By 2019, the opioid and fentanyl death rates were 44% and 50% greater in the legalizing than non-legalizing jurisdictions, respectively. When the COVID-19 pandemic hit in 2020, jurisdictions that implemented recreational marijuana legalization before 2019 had significantly greater increases in both overall opioid and fentanyl death rates than jurisdictions with medicinal-only legalization. For all opioids, the mean (95% confidence interval [CI]) 2019-to-2020 increases were 46.5% (95% CI, 36.6% to 56.3%) and 29.1% (95% CI 20.2% to 37.9%), respectively (p = 0.02). For fentanyl, they were 115.6% (95% CI, 80.2% to 151.6%) and 55.4% (95% CI, 31.6% to 79.2%), respectively (p = 0.01). Marijuana legalization is correlated with worsening of the U.S. opioid epidemic, and especially during the COVID-19 pandemic with recreational legalization.

## Introduction

As of 2022, most states and the District of Columbia (D.C.) [jurisdictions] in the U.S. had legalized medicinal marijuana (cannabis),^1^ including 20 jurisdictions that legalized recreational use, 12 of which did so during 2019-2022.^2^ The pressure for the federal government to remove its prohibition of marijuana sales and use and to leave their regulation to state law-makers is increasing, including “moving past the *gateway hypothesis* of drug use”.^3^ In October, 2022, the President pardoned federal convictions for simple marijuana possession offenses.^4^ By 2026, sales of legal recreational cannabis the U.S. are predicted to reach 37 billion dollars^5^ and the potential economic benefit has incentivized jurisdiction legalization.^6^

The U.S. also has the world’s 2nd highest cannabis-use-disorder prevalence and, by far, the world’s highest opioid death rate.^7^ To assess whether or not these two grim statistics are related, as considered by others,^8,9^ we previously compared the opioid death rate in the composite aggregate of legalizing jurisdictions with that in non-legalizing jurisdictions.^7^ With then too few evaluable recreational-legalizing jurisdictions, we did not assess them for differences in the impact of recreational versus medicinal legalization. In this study, we update all 50 states and D.C. during the last decade (2010-2019) by comparing opioid mortality rates in jurisdictions states that had or had not by start of 2019 legalized marijuana for all opioids and the fentanyl group of synthetic opioids. Because, as described in the Methods section, the COVID-19 pandemic distinctly changed prior opioid mortality trends, we also analyzed the first calendar year of the COVID-19 pandemic that began in 2020 with the latest available data and compared it with the prior calendar year, 2019.

## Methods

### Data Sources and Analytic Methods

Age-adjusted opioid death data in the U.S. were obtained from CDC WONDER.^10^ For multiple calendar-year analysis, trends were determined with Joinpoint Regression Program version 4.9.1.0,^11^ applying weighted least squares, logarithmic transformation, and standard errors provided by the Program. The Joinpoint Regression Program identifies when a trend changes to another trend, the average annual percent change (AAPC) and p-values for each trend detected, and relative comparison of concomitant trends via pairwise comparison with either parallel or non-parallel methodology, for which we selected the latter. T-tests used for comparisons assumed unequal variances.

### International Classification of Disease (ICD)

ICD codes for accidental poisoning (X40-X44), intentional self-poisoning (X60-X64), and other poisoning (Y10-Y14) were used in conjunction with following opioid T-Codes: T40.0 opium, T40.1 heroin, T40.2 other opioids, T40.3 methadone, T40.4 fentanyl and its semisynthetic derivatives, T40.6 other synthetic narcotics. These categories include morphine, hydromorphone, oxycodone, fentanyl, the semisynthetic fentanyl moieties, heroin, opium, codeine, meperidine, methadone, propoxyphene, tramadol, and other/unspecified narcotics. Because of the dramatic increase in fentanyl deaths such that by 2017 accounted for the majority of opioid deaths (Fig. 1), this category (T40.4) was also separately analyzed and referred to a *potent fentanyl*, inclusive of the more potent semisynthetic derivatives.

**Figure 1.**
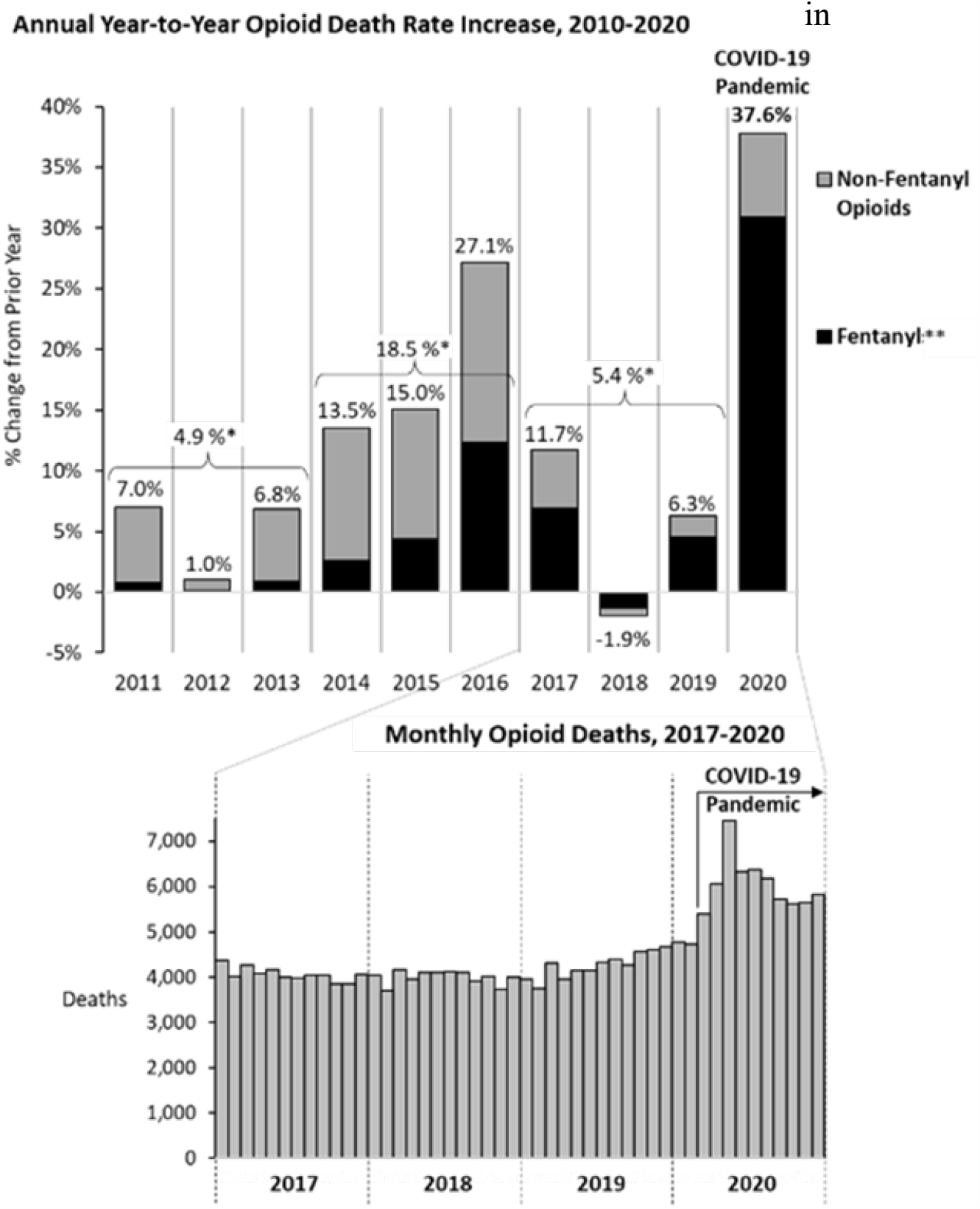
Annual Change (from year before) in Opioid Death Rate, 2010-2020, U.S., and by Portion due to Fentanyl” (black zones). Labeled percentages are increases in all opioid deaths from year before. * Percentages above horizontal brackets are 3-year averages. ** Includes semisynthetic fentanyls <u>Data Source</u>: CDC WONDER^10^

### Pre- and Intra Pandemic COVID-19 Analyses

With the onset of the COVID-19 pandemic in 2020, the opioid mortality trends in the U.S. distinctly accelerated (Fig. 1). The opioid death rate increased 37.6% from 2019 to 2020, 7 times more than the average of the prior 3 years, 5.4% (Fig. 1 upper panel). Every month in 2020 after the pandemic began in March was the highest ever recorded (Fig. 1 lower panel). The 2019 to 2020 acceleration occurred in all 50 state and D.C. jurisdictions except New Hampshire. Hence, we compared the 1^st^ year of the pandemic, 2020, with the prior year.

### Jurisdiction Classification

Supplemental Table S1 lists each state and D.C. by whether and when marijuana legalization for medicinal or recreational use was implemented, and the reference sources.^12,13,14,15,16,17,18,19,20,21,22^ As of the start of 2019, 28 jurisdictions (27 states and D.C.) had implemented marijuana legalization and 23 states that had not, as shown at the top of Figures 2 and 3 and listed in Supplemental Table S1. The legalizing group mortality trend was determined via cumulative aggregate, beginning in 2020 with those that had already implemented legalization and adding the legalizing jurisdiction thereafter during the year it implemented legalization. Georgia, North Carolina, South Carolina, Texas and Wisconsin were included in the non-legalizing group since they legalized only CBD oil for medicinal use and primarily for epilepsy. Arkansas was not included in the legalizing group since medicinal licenses were not statewide until 2020 (Supplemental Table S1).

**Figure 2.**
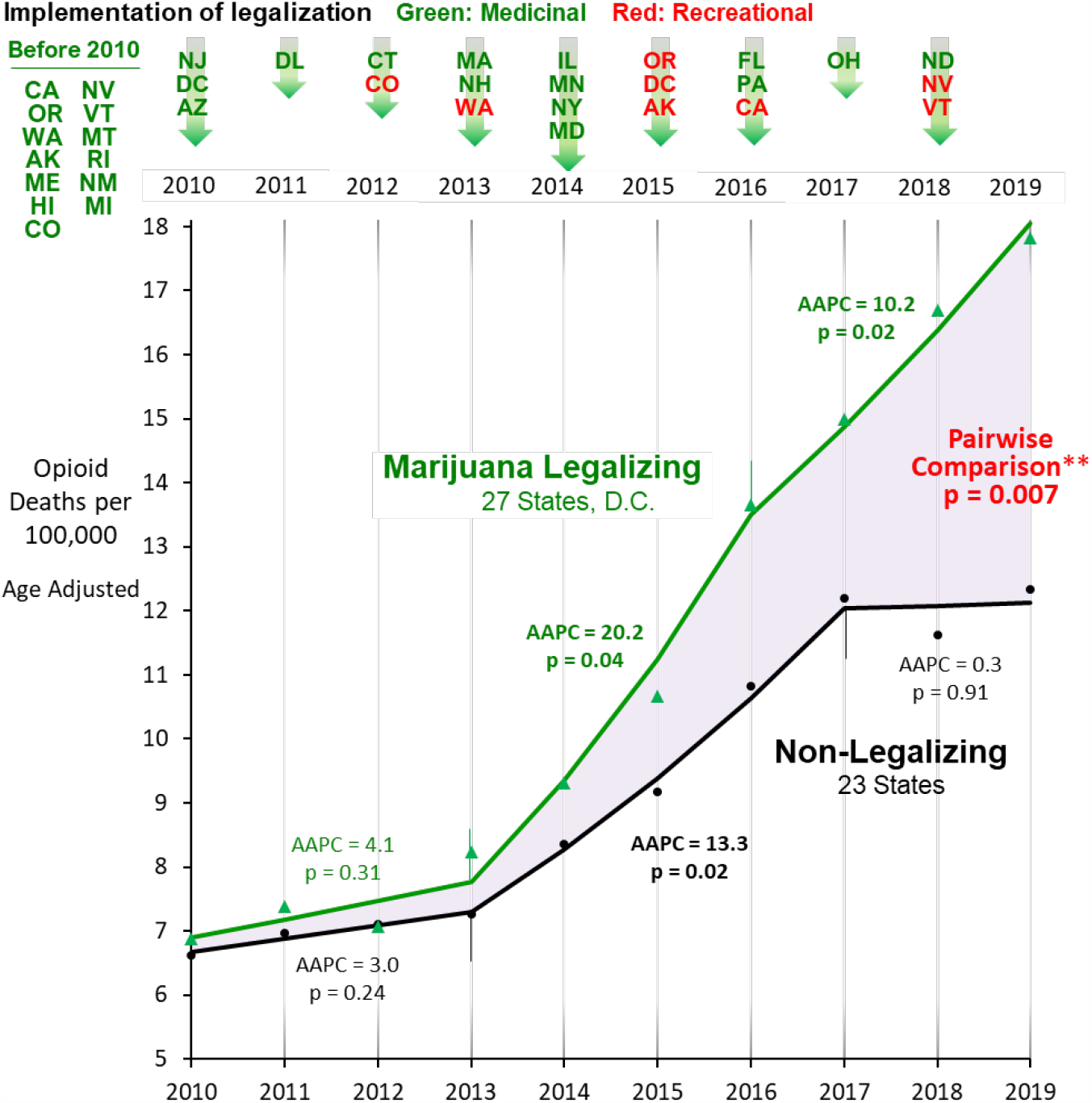
Joinpoint/AAPC* Analysis of Annual Opioid Death Rates, 2010-2020, U.S., of Non-Legalizing Jurisdictions (black data) and Cumulative Aggregate of Legalizing Jurisdictions as of 2019 (green data) * AAPC - average annual percent change ** Joinpoint non-parallel pairwise comparison analysis <u>Data Source</u>: CDC WONDER^10^and updated from Bleyer A, Barnes B, Finn K^7^

**Figure 3.**
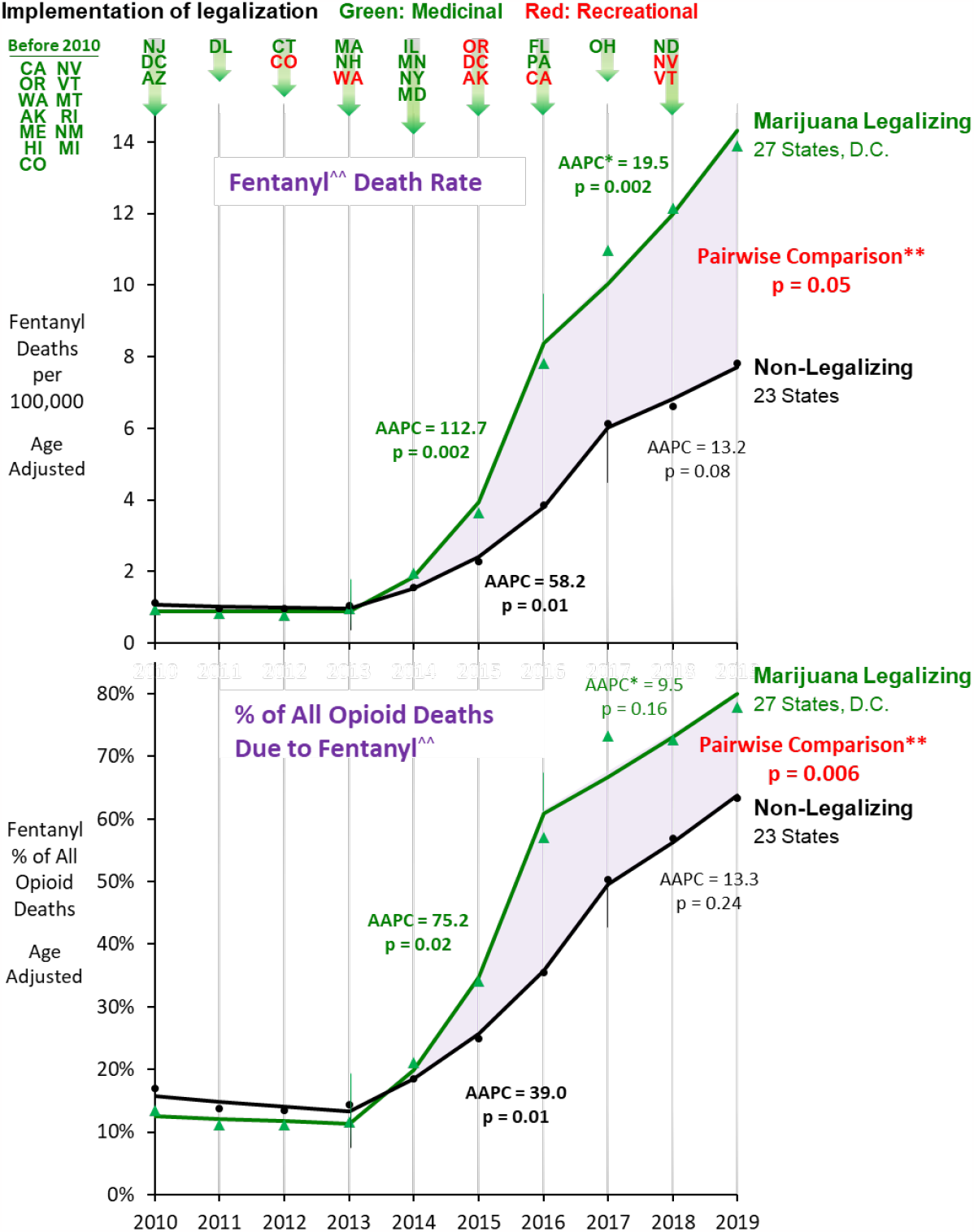
Joinpoint/AAPC* Analysis of Pre-Pandemic Annual Mean Fentanyls Death Rate (upper panel) and % of All Opioid Deaths due to Fentanyl (lower panel), 2010-2019, U.S., Non-Legalizing Jurisdictions (black data) vs. Aggregate of Legalizing Jurisdictions as of 2019 (green data). * AAPC - average annual percent change ** Joinpoint non-parallel pairwise comparison analysis ^^^^ Includes semisynthetic fentanyls <u>Data Source</u>: CDC WONDER^10^

For recreational legalization analysis, the 7 states and D.C. that implemented recreational use before 2019 were compared with 20 states that had implemented medicinal, but not recreational, use. Three states that implemented recreational use during 2019-2020, Michigan, Massachusetts and Maine, were considered within the medicinal legalization group since their recreational legalization implementation occurred too recently to expect a recreational-legalization-specific effect, especially with the dramatic increase in all but one jurisdiction during the first year of the pandemic. A sub-analysis that excluded these 3 states from the medicinal group was also conducted. Similarly, West Virginia, which implemented medicinal legalization during 2019-2020, was primarily excluded from the legalizing jurisdictions groups but also sub-analyzed as in it. Using year of legalization enactment itself instead of the year of implementation of legalization accentuated the difference in overall opioid results and did not significantly alter the fentanyl results.^7^ Difference-in-difference methodology was unnecessary to compare subsequent trends since the rates in the two groups were nearly identical for the initial three years of comparison.

## Results

### Pre-pandemic Trends

The opioid death rate in the U.S. began to increase at a statistically-significant rate in 2014 (Fig. 2), after jurisdictions began to legalize recreational use of marijuana, with Colorado and Washington as the first two states, in 2012-2013. Thereafter, the opioid death rate increased significantly more rapidly in legalizing jurisdictions, at an AAPC of 20.2 (p = 0.04) during 2013-2016 and 10.2 (p = 0.02) during 2016-2019, in contrast to an AAPC of 15.3 (p = 0.02) during 2013-2017 and no increase during 2017-2019 (AAPC = 0.3, p = 0.91) in non-legalizing jurisdictions (Fig. 2). By 2019, the opioid death rate was 44% greater in the legalizing than non-legalizing group. Over the 2010-2019 decade, pairwise comparison was highly statistically significant (p = 0.007) (Fig. 2).

The potent fentanyl death rate also increased after recreational use of marijuana began during 2012-2013, especially in legalizing jurisdictions (Fig. 3). During 2013-2016, the increase was twice as fast in legalizing than non-legalizing jurisdictions, and during 2017-2019 it continued to rise at a rate that was statistically significant (p = 0.002) whereas it slowed to a non-statistically significant rate increase in non-legalizing jurisdictions (p = NS) (Fig. 3 upper panel). By 2019, the potent fentanyl death rate was 50% greater in the legalizing than non-legalizing jurisdictions, at 13.9 and 7.8 per 100,000 respectively, and the total increase during 2000-2019 was greater in the legalizing jurisdictions (pairwise comparison p = 0.05) (Fig. 3 upper panel). During 2010-2019, there were a greater percentage of opioid deaths due to fentanyl (pairwise comparison p = 0.006) (Fig. 3 lower panel). By 2019, the proportion of opioid deaths due to potent fentanyl was 80% in legalizing jurisdictions and 63% in non-legalizing jurisdictions (Fig. 3 lower panel).

### First-Year Pandemic Trends

When the COVID-19 pandemic distinctly accelerated the opioid death rate, the 8 jurisdictions that implemented legalization of recreational marijuana before 2019 (Fig. 4 red data) had, as a group, a 30.4 absolute % greater increase than the 20 jurisdictions that implemented medicinal but not recreational legalization (57.7% vs. 27.3%) (Fig. 4 green data). At the jurisdiction level, the opioid death rate was greater in recreational-legalizing than medicinally-legalizing jurisdictions (T = 2.22, p = 0.02), with mean (95% C.I.) increases of 46.5% (36.6%, 56.3%) and 29.1% (20.2%, 37.9%), respectively (Fig. 4).

**Figure 4.**
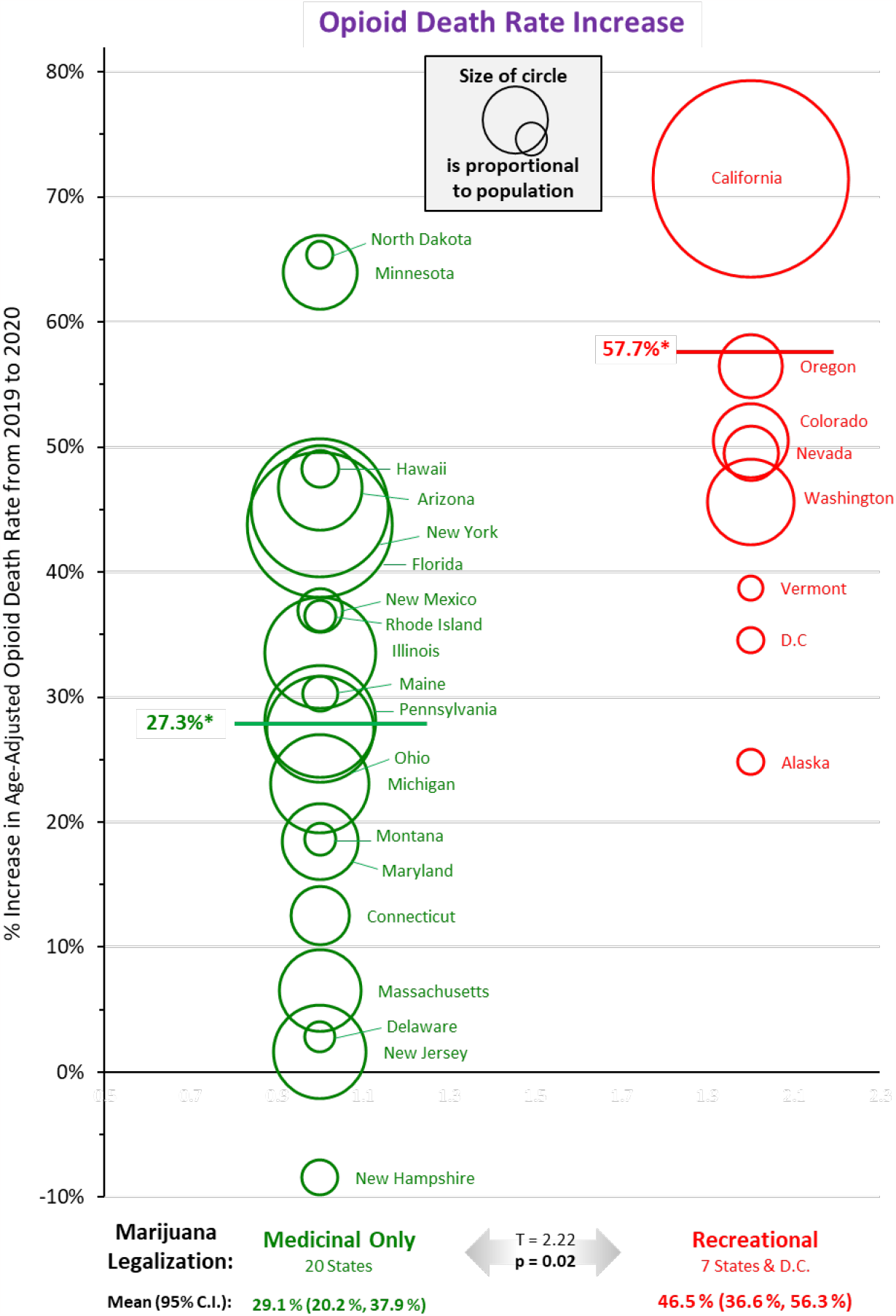
Increase in Opioids Death Rate from 2019 to First Year of Pandemic, 2020, by Recreational and Medicinal Only Legalizing Jurisdictions. * Group mean (jurisdictions combined) <u>Data Source</u>: CDC WONDER^10^

For potent fentanyl, the death rate in the recreational legalization jurisdictions accelerated at an 87.1 absolute % greater increase than in jurisdictions that implemented medicinal but not recreational legalization (123.3% vs. 36.2%) (Fig. 5). At the jurisdiction level, the opioid death rate was also greater (T = 2.69, p = 0.01) for recreational-legalizing jurisdictions than the medicinally-legalizing jurisdictions, with mean (95% C.I.) increases of 115.6% (80.2%, 151.6%) and 55.4% (31.6%, 79.2%), respectively (Fig. 5).

**Figure 5.**
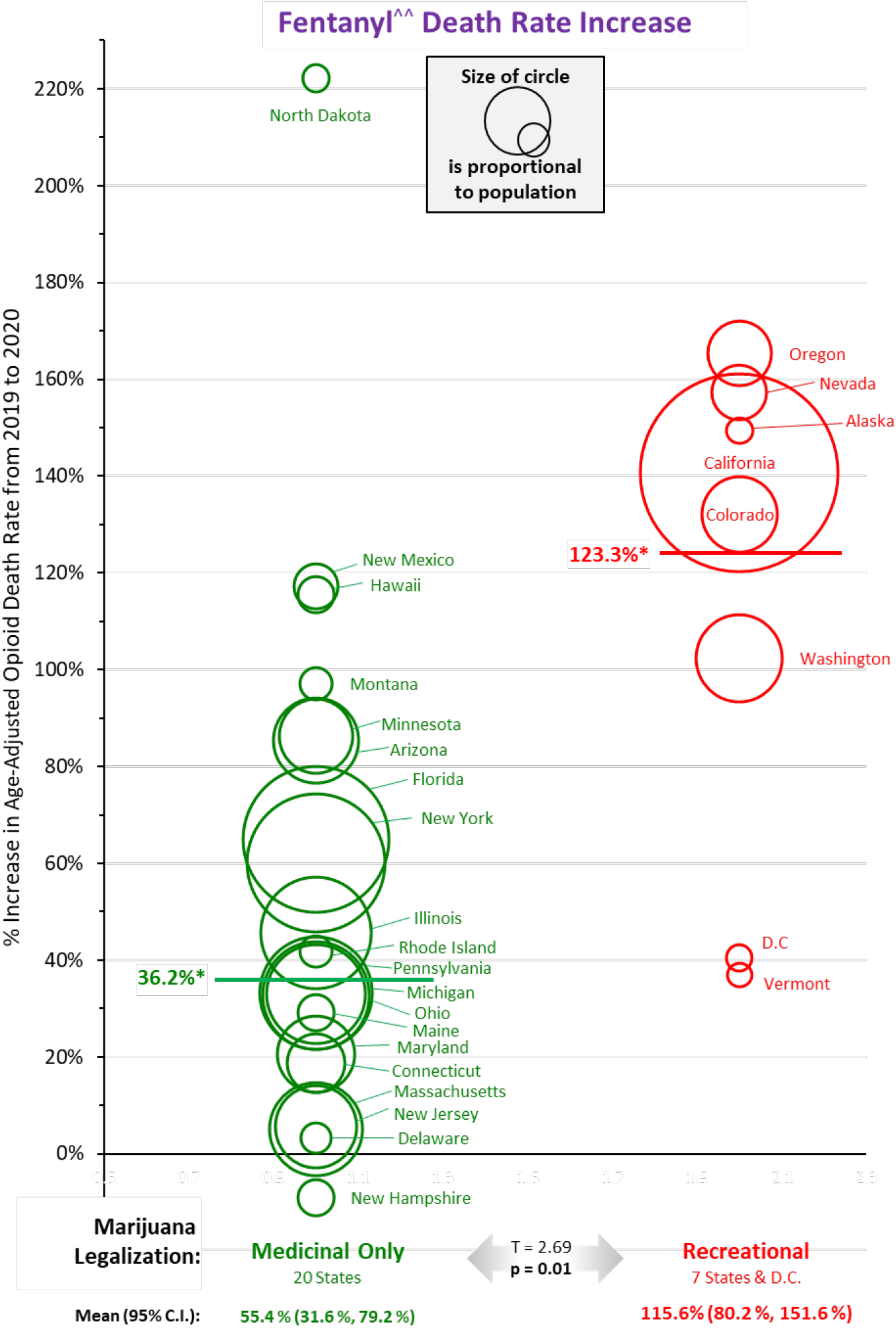
Increase in Fentanyl ^^ ^^ Death Rate from 2019 to First Year of Pandemic, 2020, by Recreational and Medicinal Only Legalizing Jurisdictions. * Group mean (jurisdictions combined) ^^ ^^ Includes semisynthetic fentanyls <u>Data Source</u>: CDC WONDER^10^

If the 3 states that began recreational legalization during 2019-2020, Maine, Massachusetts, and Michigan, are removed from the medical legalizing group, the recreational legalizing group still had a statistically-significant greater increase for both all opioids (T = 2.20, p = 0.02) and the fentanyl group of opioids (T = 2.39, p = 0.02). If the state that began medicinal legalization during 2019-2020, West Virginia, is included in the medical legalizing group, the recreational legalizing group continued to have a statistically-significant greater increase for both all opioids (T = 2.25, p = 0.02) and potent fentanyl (T = 2.74, p = 0.01). California is the state with the largest population and greatest increase in opioid death rate. Excluding it from the recreational legalization group did not eliminate the statistically-significant greater opioid death rate increase in the recreational than medical legalizing jurisdictions for either opioids in general (T = 2.27, p = 0.02) or the fentanyl group of opioids (T = 2.38, p = 0.02).

## Discussion

As analyzed, U.S. state and D.C. data suggest that marijuana legalization has been associated with worsening of opioid mortality, whether primarily due to conventional opioids or to the fentanyl group of opioids. The most recent trend indicates that recreational-use legalization is associated with a greater opioid mortality increase. If so, jurisdictions that have legalized marijuana have contributed disproportionately to the country’s opioid crisis.

### Evidence for Biologic Mechanism of Marijuana Legalization Increasing Opioid Mortality

That marijuana legalization may lead to opioid use is biologically plausible via a *gateway* mechanism since cannabinoids act in part via opioid receptors.^23^ Cannabinoids also increase dopamine concentrations.^24,25^

Clinical evidence for gateway sequencing has been reported in multiple studies. In the U.S., 9% of adults progressed to other illicit drug use during the second year after first marijuana use, 36% within a decade, and nearly half thereafter.^26^ Non-medical prescription opioid use and opioid use disorder increased nearly 6- and 8-fold, respectively within 3 years of marijuana use.^27^ Medical marijuana use was positively associated with greater use and misuse of prescription opioids.^28^ Self-reported marijuana use during injury recovery was associated with a subsequent increased amount and duration of opioid use.^29^ In New York State, opioid use was found to approximately double on days when marijuana was used.^30^ In a national database of prescription claims, opioid-naïve patients who were marijuana users undergoing lumbar spinal fusions had a higher risk of developing post-operative opioid dependence than non-marijuana users, despite having decreased daily dosages of opioids.^31^ A similar pattern was observed In Boston.^32^ Among college students, marijuana users were 12 times more likely to use opioids than non-users and higher levels of marijuana use was associated with greater likelihood of using opioids.^33^ Among Maryland students followed for 12 years after elementary school, marijuana at age 14 was associated with more frequent use of opioids at age 19.^34^ Among Los Angeles high-school students, each of five different marijuana products was associated with subsequent initiation of illicit drug use, including heroin, and of prescription opioids.^35^ Among pregnant women, the rate of opioid-related treatment admissions was 2.5 fold greater in states that legalized medicinal marijuana.^36^ Most recently, evidence from national surveys indicate that Americans with cannabis use disorder had nearly 7 times the odds of also having opioid use disorder^37^ and that after 2-3 years of recreational legalization the frequency of misusing prescription opioids increases.^38^

Other countries have had similar evidence. In Canada, marijuana use was found to lead to premature withdrawal from opioid addiction treatment programs.^39^ In Australia, a 4-year prospective-cohort study of 1,514 patients with chronic non-cancer pain, those who used marijuana daily or near-daily used more opioids than those who did not.^40^ A meta-analysis of 6 studies in Australia, New Zealand and U.S. between 1977 and 2017 indicated that transitioning from marijuana to opioid use, abuse or dependence was 2.5-2.8 times greater than in non-marijuana users.^41^

### Prior Studies of Marijuana Legalization’s Effect on Opioid Mortality

Initial reports based on a limited number of U.S. states suggested that marijuana legalization reduced opioid mortality.^13,42,43^ One of these reports was based on 2006-2011 data, before the subsequent acceleration of opioid mortality.^44^ A report in 2018 concluded that medical marijuana legalization was associated with a 30% reduction in Schedule III opioid Medicaid prescriptions.^45^ Two county-level studies in states that allowed marijuana dispensaries to operate had lower opioid-mortality rates in counties with medical and recreational dispensaries,^46,47^ albeit the county-level methodology utilized has been negatively critiqued.^7,48^

At the national level, an update of aforementioned studies by other investigators indicates that legal medical marijuana is associated with higher opioid mortality, particularly when available through retail dispensaries, and that recreational marijuana may be correlated with greater death rates relative to the counterfactual of no legal cannabis.^49^ Opioid overdose death rates during 2012-2017 in states that legalized medicinal marijuana had a greater increase in opioid mortality than in those that did not (p<0.01).^50^ In Massachusetts, Maine, Nevada and California, per-capita opioid-related emergency department visit rates initially decreased in states that legalized recreational use but began to increase within 6 months after legalization,^51^ as of the last year of data in 2017 and before most recreational-legalizing jurisdictions had an acceleration in their opioid mortality.^7^

In Colorado, three studies have not supported the initial impression of a marijuana legalization benefit on opioid mortality,^52,53,54^ one of which not only found that the original analysis did not hold over the longer period, but the association between state medical cannabis laws and opioid overdose mortality reversed direction from −21% to +23% and remained positive after accounting for recreational cannabis laws.^52^ Another had opioid-related ER visits and admissions to the Colorado Hospital Association increase 51% and 45%, respectively, during the first 4 years after recreational legalization.^54^ In Washington and Colorado, hospital admissions for opioid substance abuse did not decrease during the first 2-4 years, during 2013-2017, after recreational legalization.^55^ In Washington state, recreational marijuana legalization was not followed by a reduction in opioid compliance rate in patients treated with opioids for chronic pain during the first year after recreational legalization.^56^ As we previously reported, the opioid mortality increase in these both Colorado and Washington two states began after 2017 and more than 2 years after recreational legalization.^7^

### Limitations

The most important limitation of our study is its ecological design that does not establish causation. Factors other than marijuana legalization may have resulted in marijuana legalizing jurisdictions having a higher opioid death rate. Most notably, legalizing jurisdictions may be more substance use/abuse/dependent-oriented and thus the local culture may be responsible for both, rather than marijuana legalization exacerbating opioid mortality. Legalizing and non-legalizing jurisdictions may differ in socioeconomic status, race/ethnicity, and medical, psychosocial, and psychiatric diagnoses that may have caused more opioid deaths in legalizing jurisdictions. In that opioid use disorder is a “disease of despair” brought about by economic hardship, the economic issue is particularly concerning, albeit the 2020 gross domestic product per capita in the legalizing states we previously analyzed was greater than in the non-legalizing states.^7^ As to the greater opioid mortality increase during the pandemic in recreationally-legalizing jurisdictions, several were states that considered marijuana businesses as “essential” and allowed to remain open whereas drug treatment programs were closed and more opioid-use-disorder patients were unable to access care.

The most contradictory report relative to our analysis is a study of recreational marijuana-legalizing states of Colorado, Washington, Oregon, Alaska, Nevada, California, and Massachusetts that found they had an 11% average (range 3%-28%) reduction in opioid overdose fatalities.^57^ This study did not include 2020 when the opioid mortality increased the most and that we specifically evaluated, did not compare the opioid mortality rate in recreational states with either medicinal-only or non-legalizing states, and did not include D.C. Another study that counters our findings is a review of individual opioid prescriptions during 2011-2018 that associated recreational and medicinal cannabis access laws with fewer morphine milligram-equivalents and total days’ supply of opioids prescribed, number of patients receiving opioids, and probability a provider prescribes any opioids net of any offsetting effects.^58^

### Strengths

The current investigation also has several advantages over prior reports. It adds 9, 6, 4, 2, and 2 additional follow-up years to the prior studies.^13,42,43,44,46^, respectively Compared to the most recently reported state-level analysis^44^ that analyzed 2000-2011 data, it includes data up to 2020. A prior report also showed a reversal of initial benefit of marijuana legalization to worsening opioid mortality.^52^ Our analysis of their data shows a divergence in the opioid death rates during 2012-2017 that is similar to what we observed during those years (Fig. 2). Our study adds two more years of data and D.C., and also differs in that our control group was states that had not legalized marijuana whereas their control group began with all states and excluded those that legalized when they did. Also, we included separate analyses of the T40.4 category of fentanyl and semi-synthetic analogues and we included heroin and opium that were either not assessed in prior studies^13,54,59^ or specified.^28,43^

## Conclusions

At the jurisdiction level in the U.S., marijuana legalization during the past decade was associated with a greater acceleration in the opioid death rate. During the first year of the COVID-19 pandemic, the opioid mortality acceleration occurred significantly more in jurisdictions that legalized recreational marijuana than in those that legalized medical, but not recreational, use. A strikingly greater increase in marijuana sales during the pandemic may have contributed to an even greater increase in the country’s opioid mortality epidemic during its 2^nd^ year and, possibly, years subsequent to the pandemic.^60^

## Data Availability

All data produced in the present study are available upon reasonable request to the authors. The data sources used are available online at CDC WONDER via http://wonder.cdc.gov/mcd-icd10.html.

## Acknowledgements

The authors thank David Hemenway, PhD, Professor of Health Policy at the Harvard School of Public Health and Director of the Harvard Injury Control Research Center and the Harvard Youth Violence Prevention Center, who provided suggestions to help verify the findings and improve interpretations of the analyses.

## References

1. Victor B, Hager K, Stacy S. Re-legalizing cannabis for medical use in the USA. J Public Health (Oxf). 2022 Aug 25;44(3):679–684. doi:10.1093/pubmed/fdab066.

2. State Medical Cannabis Laws, National Conference of State Legislatures (NCSL). https://www.ncsl.org/research/health/state-medical-marijuana-laws.aspx#3.Accessed May 15, 2023.

3. Arnold JF, Sade RM. Regulating marijuana use in the United States: Moving past the gateway hypothesis of drug use. J Law Med Ethics. 2020;48(2):275–278. doi:10.1177/1073110520935339.

4. Presidential proclamation on marijuana possession. The United States Department of Justice. https://www.justice.gov/pardon/presidential-proclamation-marijuana-possession.Accessed May 15, 2023.

5. Sales of legal recreational cannabis in the United States from 2021 to 2026. https://www.statista.com/statistics/933384/legal-cannabis-sales-forecast-us/.Accessed May 15, 2023.

6. Dills A, Goffard S, Miron J, Partin E. The effect of state marijuana legalizations: 2021 Update. 2021(Feb 2), https://www.cato.org/policy-analysis/effect-state-marijuana-legalizations-2021-update. Accessed May 18, 2023.

7. Bleyer A, Barnes B, Finn K. United States marijuana legalization and opioid mortality epidemic during 2010-2020 and pandemic implications. J Natl Med Association 2022;114(4):412–425. Online 22 April 2022. doi.org/10.1016/j.jnma.2022.03.004.

8. Kaufman DE, Nihal AM, Leppo JD, Staples KM, McCall KL, Piper BJ. Opioid mortality following implementation of medical cannabis programs in the United States. Pharmacopsychiatry. 2021 Mar;54(2):91–95. doi:10.1055/a-1353-6509. Epub 2021 Feb 23.

9. Athanassiou M, Dumais A, Zouaoui I, Potvin S. The clouded debate: A systematic review of comparative longitudinal studies examining the impact of recreational cannabis legalization on key public health outcomes. Front Psychiatry. 2023 Jan 11;13:1060656. doi:10.3389/fpsyt.2022.1060656.

10. Centers for Disease Control and Prevention, National Center for Health Statistics. Multiple Cause of Death 1999-2020 on CDC WONDER Online Database, released in 2021. Data are from the Multiple Cause of Death Files, 1999-2020, as compiled from data provided by the 57 vital statistics jurisdictions through the Vital Statistics Cooperative Program. Accessed at http://wonder.cdc.gov/mcd-icd10.html on May 15, 2023.

11. Joinpoint Regression Program, Version 4.9.0.0. March, 2021; Statistical Research and Applications Branch, National Cancer Institute. Statistical Research and Applications Branch, National Cancer Institute. Kim HJ, Fay MP, Feuer EJ, Midthune DN. Permutation tests for joinpoint regression with applications to cancer rates. Stat Med. 2000;19:335–351 (correction: 2001;20:655).

12. State Medical Cannabis Laws, National Conference of State Legislatures (NCSL). http://www.ncsl.org/research/health/state-medical-marijuana-laws.aspx#3.Accessed May 15, 2023.

13. Powell D, Pacula RL, Jacobson M. Do medical marijuana laws reduce addictions and deaths related to pain killers? J Health Econ. 2018;58 (March):29–42. doi:10.1016/j.jhealeco.2017.12.007.

14. Martins SS, Segura LE, Levy NS, et al. Racial and ethnic differences in cannabis use following legalization in US states with medical cannabis laws. JAMA Netw Open. 2021;4(9):e2127002. doi:10.1001/jamanetworkopen.2021.27002.

15. Office of Cannabis Policy. State of Maine Office of Administrative and Financial Services. https://www.maine.gov/dafs/omp/adult-use.Accessed May 15, 2023.

16. 16. Cannabis in Michigan. https://en.wikipedia.org/wiki/Cannabis_in_Michigan. Accessed May 15, 2023.

17. 17. Cannabis in Massachusetts. https://en.wikipedia.org/wiki/Cannabis_in_Massachusetts. Accessed May 15, 2023.

18. Low THC Oil Registry. Georgia Department of Public Health. https://dph.georgia.gov/low-thc-oil-registry. Accessed May 15, 2023.

19. 19. Cannabis in Georgia. https://en.wikipedia.org/wiki/Cannabis_in_Georgia_(U.S._state). Accessed May 15, 2023.

20. 20. Cannabis in Louisiana. https://en.wikipedia.org/wiki/Cannabis_in_Louisiana. Accessed May 15, 2023.

21. 21. Cannabis in [State name]. https://en.wikipedia.org/wiki/Cannabis_in_[state name]. 2023, Accessed May 15, 2023.

22. Regulating Medical Marijuana. Louisiana Department of Health. https://ldh.la.gov/assets/oph/Rulemaking/noi/Medical_Marijuana_NOI_cv_Register.pdf. Accessed May 15, 2023.

23. Scavone JL, Sterling RC, Van Bockstaele EJ. Cannabinoid and opioid interactions: implications for opiate dependence and withdrawal. Neuroscience. 2013;248:637–654. doi.org/10.1016/j.neuroscience.2013.04.034.

24. Tanda G, Pontieri FE, Di Chiara G. Cannabinoid and heroin activation of mesolimbic dopamine transmission by a common μ1 opioid receptor mechanism. Science. 1997;276(5321):2048–2050.

25. Ashton CH. Pharmacology and effects of cannabis: a brief review. Br J Psychiatry. 2001;178:101–106. doi.org/10.1192/bjp.178.2.101.

26. Secades-Villa R, Garcia-Rodríguez O, Jin CJ, Wang S, Blanco C. Probability and predictors of the cannabis gateway effect: a national study. Int J Drug Policy. 2015 Feb;26(2):135–42. doi:10.1016/j.drugpo.2014.07.011.

27. Olfson M, Wall MM, Liu SM, Blanco C. Cannabis use and risk of prescription opioid use disorder in the United States. Am J Psychiatry. 2018 Jan 1;175(1):47–53. doi.org/10.1176/appi.ajp.2017.17040413.

28. Caputi TL, Humphreys K. Medical marijuana users are more likely to use prescription drugs medically and non-medically. J Addict Med. 2018;12(4):295–299. doi.org/10.1097/ADM.

29. Bhashyam AR, Heng M, Harris MB, Vrahas MS, Weaver MJ. Self-reported marijuana use is associated with increased use of prescription opioids following traumatic musculoskeletal injury. J Bone Joint Surg. 2018 (Dec 19);100 (24):2095–2102.

30. Gorfinkel LR, Stohl M, Greenstein E, Aharonovich E, Olfson M, Hasin D. Is cannabis being used as a substitute for non-medical opioids by adults with problem substance use in the United States? A within-person analysis. Addiction. 2021 May;116(5):1113–1121. doi:10.1111/add.15228.

31. Khalid SI, Jiang S, Khilwani H, Thomson K, Mirpuri P, Mehta AI. Postoperative opioid use among opioid-naïve cannabis users following single-level lumbar fusions. World Neurosurg. 2023 Apr 6:S1878-8750(23)00475-8. doi:10.1016/j.wneu.2023.04.001. Epub ahead of print.

32. Moon AS, LeRoy TE, Yacoubian V, Gedman M, Aidlen JP, Rogerson A. Cannabis use is associated with increased use of prescription opioids following posterior lumbar spinal fusion surgery. Global Spine J. 2022 May 10:21925682221099857. doi:10.1177/21925682221099857.

33. Palfai TP, Tahaney KD, Winter MR. Is marijuana use associated with health promotion behaviors among college students? Health-promoting and health-risk behaviors among students identified through screening in a university student health services center. J Drug Issues. 2016;46:41–50.

34. Thrul J, Rabinowitz JA, Reboussin BA, Maher BS, Ialongo NS. Adolescent cannabis and tobacco use are associated with opioid use in young adulthood 12-year longitudinal study in an urban cohort. Addiction. 2021 Mar;116(3):643–650. doi:10.1111/add.15183.

35. Braymiller JL, Riehm KE, Meier M, et al. Associations of alternative cannabis product use and poly-use with subsequent illicit drug use initiation during adolescence. Psychopharmacology (Berl). 2023 Mar 3. Epub ahead of print. doi:10.1007/s00213-023-06330-w.

36. Meinhofer A, Witman A, Murphy SM, Bao Y. Medical marijuana laws are associated with increases in substance use treatment admissions by pregnant women. Addiction. 2019;114(9):1593–1601. doi.org/10.1111/add.14661.

37. Harton MR, Adams ZW, Parker MA. Associations of alcohol use disorder, cannabis use disorder, and nicotine dependence with concurrent opioid use disorder in U.S. adults. Exp Clin Psychopharmacol. 2023 May 11. doi:10.1037/pha0000649. Epub ahead of print.

38. Ali MM, McClellan C, Mutter R, Rees DI. Recreational marijuana laws and the misuse of prescription opioids: Evidence from National Survey on Drug Use and Health microdata. Health Econ. 2023 Feb;32(2):277–301. doi:10.1002/hec.4620. Epub 2022 Nov 5.

39. Franklyn AM, Eibl JK, Gauthier GJ, Marsh DC. The impact of cannabis use on patients enrolled in opioid agonist therapy in Ontario, Canada. PLoS One. 2017 Nov 8;12(11):e0187633. doi:10.1371/journal.pone.0187633.

40. Campbell G, Hall WD, Peacock A, Lintzeris N, Bruno R, Larance B, Nielsen S, Cohen M, Chan G, Mattick RP, Blyth F Shanahan M, Dobbins T, Farrell M, Degenhardt L. Effect of cannabis use in people with chronic non-cancer pain prescribed opioids: findings from a 4-year prospective cohort study. Lancet Public Health. 2018 (July);3:e341–350.

41. Wilson J, Mills K, Freeman TP, Sunderland M, Visontay R, Marel C. Weeding out the truth: a systematic review and meta-analysis on the transition from cannabis use to opioid use and opioid use disorders, abuse or dependence. Addiction. 2022;117(2):284–298. doi:10.1111/add.15581.

42. Bachhuber MA, Saloner B, Cunningham CO, Barry CL. Medical cannabis laws and opioid analgesic overdose mortality in the United States, 1999-2010. JAMA Intern Med. 2014;174(10):1668–1673. doi:10.1001/jamainternmed.2014.4005.

43. Livingston MD, Barnett TE, Delcher C, Wagenaar AC. Recreational cannabis legalization and opioid-related deaths in Colorado, 2000-2015. Amer J Public Health. 2017;107(11):1827–1829.

44. Kim JH, Martins SS, Shmulewitz D, Hasin D. Association between fatal opioid overdose and state medical cannabis laws in US national survey data, 2000-2011. International Journal of Drug Policy. 2022 (January); 99: 103449, ISSN 0955-3959. doi.org/10.1016/j.drugpo.2021.103449.

45. Liang D, Bao Y, Wallace M, Grant I, Shi Y. Medical cannabis legalization and opioid prescriptions: evidence on US Medicaid enrollees during 1993-2014. Addiction. 2018 Nov;113(11):2060–2070. doi:10.1111/add.14382

46. Hsu G, Kovács B. Association between county level cannabis dispensary counts and opioid related mortality rates in the United States: panel data study. BMJ. 2021;372:m4957. doi.org/10.1136/bmj.m4957.

47. Castillo-Carniglia A, Rivera-Aguirre A, Santaella-Tenorio J, et al. Changes in opioid and benzodiazepine poisoning deaths after cannabis legalization in the US: A county-level analysis, 2002-2020. Epidemiology. 2023 Mar 16. doi:10.1097/EDE.0000000000001609.

48. Monnat SM, Peters DJ, Berg MT, Hochstetler A. Using census data to understand county-level differences in overall drug mortality and opioid-related mortality by opioid type. American Journal of Public Health, 2019;109:1084–1091. doi.org/10.2105/AJPH.2019.305136.

49. Mathur NK, Ruhm CJ. Marijuana legalization and opioid deaths. J Health Econ. 2023 Mar;88:102728. Epub 2023 Jan 6 doi:10.1016/j.jhealeco.2023.102728]

50. Kaufman DE, Nihal AM, Leppo JD, Staples KM, McCall KL, Piper BJ. Opioid mortality following implementation of medical cannabis programs in the United States. Pharmacopsychiatry. 2021 Mar;54(2):91–95. doi:10.1055/a-1353-6509. Epub 2021 Feb 23.

51. Drake C, Wen J, Hinde J, Wen H. Recreational cannabis laws and opioid-related emergency department visit rates. Health Econ. 2021;30:2595–2605. doi:10.1002/hec.4377.

52. Shover CL, Davis CS, Gordon SC, Humphreys K. Association between medical cannabis laws and opioid overdose mortality has reversed over time. Proc Natl Acad Sci U S A. 2019;116(26):12624–12626.

53. Alcocer JJ. Exploring the effect of Colorado’s recreational marijuana policy on opioid overdose rates, Public Health. 2020;185;8–14. doi.org/10.1016/j.puhe.2020.04.007.

54. Wang GS, Buttorff C, Wilks A, et al. Comparison of hospital claims and poison center data to evaluate health impact of opioids, cannabis and synthetic cannabinoids. Am J Emerg Med. 2022 Mar;53:150–153. doi:10.1016/j.ajem.2022.01.004. Epub 2022 Jan 11.

55. Mennis J, Stahler GJ. Adolescent treatment admissions for marijuana following recreational legalization in Colorado and Washington. Drug Alcohol Depend. 2020 May 1;210:107960. doi:10.1016/j.drugalcdep.2020.107960. Epub 2020 Mar 19.

56. Lo SY, Winston-McPherson GN, Starosta AJ, Sullivan MD, Baird GS, Hoofnagle AN, Greene DN. Cannabis legalization does not influence patient compliance with opioid therapy. Am J Med. 2019 Mar;132(3):347–353. doi:10.1016/j.amjmed.2018.11.002.

57. Marinello S, Powell LM. The impact of recreational cannabis markets on motor vehicle accident, suicide, and opioid overdose fatalities. Soc Sci Med. 2023 Mar;320:115680. doi:10.1016/j.socscimed.2023.115680.

58. McMichael BJ, Van Horn RL, Viscusi WK, The impact of cannabis access laws on opioid prescribing, J Health Economics. 2020:69,102273. 10.1016/j.jhealeco.2019.102273.

59. Segura LE, Mauro CM, Levy NS, Khauli N, Philbin MM, Mauro PM, Martins SS. Association of US medical marijuana laws with nonmedical prescription opioid use and prescription opioid use disorder. JAMA Netw Open. 2019;2(7):e197216. Published 2019 Jul 3. doi.org/10.1001/jamanetworkopen.2019.7216.

60. Schauer GL, Dilley JA, Roehler DR, et al. Cannabis sales increases during COVID-19: Findings from Alaska, Colorado, Oregon, and Washington. Int J Drug Policy. 2021 Dec;98:103384. doi:10.1016/j.drugpo.2021.103384. Epub 2021 Aug 4.

