## Supplemental Table S1 for "United States Marijuana Legalization and Opioid Mortality Trends Before and During the First Year of the COVID-19 Pandemic"

**State and District of Columbia Marijuana Legalization Implementation Status as of January 1, 2019**

| Jurisdiction | Legalization Implementation Year | | | | Comments and References |
| --- | --- | --- | --- | --- | --- |
|  | Medicinal | Recreational | | |  |
| **Marijuana Legalization Implemented before 2019:**  **20 Jurisdictions of which 8 also implemented recreational legalization before January 1, 2019** | | | | | |
| [California](https://en.wikipedia.org/wiki/California) | 1996 ^†^ | 2017 ^††^ | | Medicinal legalization was implemented >2 years before 1^st^ available opioid mortality data. | |
| [Oregon](https://en.wikipedia.org/wiki/Oregon) | 1998 ^†^ | 2015 ^††^ | |  | |
| [Washington](https://en.wikipedia.org/wiki/Washington_(state)) | 1998 ^†^ | 2013 ^††^ | |  | |
| [Alaska](https://en.wikipedia.org/wiki/Alaska) | 1999 ^†^ | 2015 ^††^ | |  | |
| [Maine](https://en.wikipedia.org/wiki/Maine) | 1999 ^†^ | 2020 | | Governor veto of recreational legalization overturned in May 2018, but sales delayed until 2020 (Office of Cannabis Policy, State of Maine, 2023). | |
| [Hawaii](https://en.wikipedia.org/wiki/Hawaii) | 2000 ^†^ |  | |  | |
| [Nevada](https://en.wikipedia.org/wiki/Nevada) | 2001 ^†^ | 2017 ^††^ | |  | |
| [Colorado](https://en.wikipedia.org/wiki/Colorado) | 2003 ^†^ | 2012 ^††^ | |  | |
| [Vermont](https://en.wikipedia.org/wiki/Vermont) | 2004 ^†^ | 2018 | |  | |
| [Montana](https://en.wikipedia.org/wiki/Montana) | 2004 ^†^ |  | |  | |
| [Rhode Island](https://en.wikipedia.org/wiki/Rhode_Island) | 2006 ^†^ |  | |  | |
| [New Mexico](https://en.wikipedia.org/wiki/New_Mexico) | 2007 ^†^ |  | |  | |
| [Michigan](https://en.wikipedia.org/wiki/Michigan) | 2009 ^†,††^ | 2019 | | Recreational use approved by Michigan voters in November 2018 and implemented during 2019-2020 (Cannabis in Michigan, 2023). | |
| [New Jersey](https://en.wikipedia.org/wiki/New_Jersey) | 2010 ^†,††^ |  | |  | |
| [D.C.](https://en.wikipedia.org/wiki/Washington,_D.C.) | 2010 ^†,††^ | 2015 ^††^ | |  | |
| [Arizona](https://en.wikipedia.org/wiki/Arizona) | 2010 ^†,††^ |  | |  | |
| [Delaware](https://en.wikipedia.org/wiki/Delaware) | 2011 ^†,††^ |  | |  | |
| [Connecticut](https://en.wikipedia.org/wiki/Connecticut) | 2012 ^†,††^ |  | |  | |
| [Massachusetts](https://en.wikipedia.org/wiki/Massachusetts) | 2013 ^†,††^ | 2019 | | Recreational use legalized in 2017 but implementation delayed by Governor with signed legislation extending the start date for legal licensed recreational cannabis until March 2019; 1^st^ 2 stores opened on November 20, 2018 (Cannabis in Massachusetts, 2023) | |
| [New Hampshire](https://en.wikipedia.org/wiki/New_Hampshire) | 2013 ^†,††^ |  | |  | |
| [Illinois](https://en.wikipedia.org/wiki/Illinois) | 2014 ^†,††^ |  | |  | |
| [Minnesota](https://en.wikipedia.org/wiki/Minnesota) | 2014 ^†,††^ |  |  | | |
| [New York](https://en.wikipedia.org/wiki/New_York_(state)) | 2014 ^†,††^ |  |  | | |
| [Maryland](https://en.wikipedia.org/wiki/Maryland) | 2014 ^††^ |  |  | | |
| Florida | 2016 ^††^ |  |  | | |
| [Pennsylvania](https://en.wikipedia.org/wiki/Pennsylvania) | 2018 ^††,†††^ |  | Legalized 4/17/2016 and implemented in 2017-2018 ^*^ | | |
| [Ohio](https://en.wikipedia.org/wiki/Ohio) | 2018 ^††,†††^ |  | [Legalized 6/8/2016 and implemented in 2017-2018 ^*^](https://en.wikipedia.org/wiki/Cannabis_in_Ohio) | | |
| North Dakota | 2018 ^††,†††^ |  | [2017 medicinal legislation partially enabled in May 2018 ^*^](https://en.wikipedia.org/wiki/Cannabis_in_Ohio) | | |

continued …

Supplemental Table S1	**State and District of Columbia Marijuana Legalization Implementation Status**

Continued	**as of January 1, 2019**

| **Marijuana Legalization not Implemented before 2019 (23 Jurisdictions)** | | |
| --- | --- | --- |
| [Alabama](https://en.wikipedia.org/wiki/Alabama) | ††† | Medical legalization in 2021. |
| [Arkansas](https://en.wikipedia.org/wiki/Arkansas) | ††† | [Licensed medicinal sales did not began until May 2019, at a single location (Hot Springs); its medical implementation was regarded to be in 2020.](https://en.wikipedia.org/wiki/Cannabis_in_Arkansas)* |
| [Georgia](https://en.wikipedia.org/wiki/Georgia_(U.S._state)) |  | Possession of <20 oz of low-concentration THC cannabidiol oil for medicinal use  legalized in 2015 and does not authorize physicians to prescribe (State Medical Cannabis Laws, 2023, Low THC Oil Registry, Georgia Department of Public Health, 2023, Cannabis in Georgia, 2023). |
| [Idaho](https://en.wikipedia.org/wiki/Idaho) | ††† | 2021: Only hemp growth & transport legalized. |
| [Indiana](https://en.wikipedia.org/wiki/Indiana) | ††† | CBD oil legalized in 2014 for epilepsy and for any purpose in 2018; not THC and not decriminalized. |
| [Iowa](https://en.wikipedia.org/wiki/Iowa) | ††† | 2019: Only hemp growth & transport legalized. |
| [Kansas](https://en.wikipedia.org/wiki/Kansas) | ††† | State House passed a medicinal marijuana bill 78–42 and sent the bill to the State Senate  to vote on it, which isn’t expected until 2022. |
| [Kentucky](https://en.wikipedia.org/wiki/Kentucky) | ††† | [Med bill stalled in the Senate due to the COVID-19 pandemic](https://en.wikipedia.org/wiki/COVID-19_pandemic). |
| [Louisiana](https://en.wikipedia.org/wiki/Louisiana) |  | [Medicinal use legislated in 2019 and implemented in 2020, and without patient registry ID cards (State Medical Cannabis Laws, 2023, Regulating Medical Marijuana. Louisiana Department of Health, 2023, Cannabis in Louisiana, 2023).](https://en.wikipedia.org/wiki/Cannabis_in_Louisiana) |
| [Mississippi](https://en.wikipedia.org/wiki/Mississippi) | ††† | 2011: State supreme court overruled medicinal legalization; decriminalized. |
| [Missouri](https://en.wikipedia.org/wiki/Missouri) | ††† | The first licensed sales of medical cannabis occurred on October 17, 2020. |
| [Nebraska](https://en.wikipedia.org/wiki/Nebraska) | ††† | Decriminalized. |
| [North Carolina](https://en.wikipedia.org/wiki/North_Carolina) | ††† | CBD oil only for epilepsy. |
| [Oklahoma](https://en.wikipedia.org/wiki/Oklahoma) | ††† | Decriminalized. |
| [South Carolina](https://en.wikipedia.org/wiki/South_Carolina) | ††† | CBD oil only for children with epilepsy. |
| [South Dakota](https://en.wikipedia.org/wiki/South_Dakota) | ††† | Medical legalization failed and recreational legalization nullified by State Supreme Court. |
| [Tennessee](https://en.wikipedia.org/wiki/Tennessee) | ††† | Attempt to pass medical legalization failed in 2021. |
| [Texas](https://en.wikipedia.org/wiki/Texas) | ††† | Medical use is allowed only in the form of low-THC cannabis oil, less than 1% THC  with a physician's approval and less than 0.3% THC without, approved in 2015. |
| [Utah](https://en.wikipedia.org/wiki/Utah) | ††† | On November 6, 2018, The Utah Medical Cannabis Act was passed as ballot Proposition 2. Provisions must be set by the state for dispensaries to open by January 2021. |
| [Virginia](https://en.wikipedia.org/wiki/Virginia) | ††† | As of April 2019 only 251 of the 35,404 doctors licensed to practice in Virginia  had registered with the state to write medical cannabis recommendations. |
| [Wisconsin](https://en.wikipedia.org/wiki/Wisconsin) | ††† | Legislated in 2017: CBD oil only for med purpose if a doctor has certified the oil is being used to treat a medical condition (State Medical Cannabis Laws, 2023). |
| West Virginia | ††† | [Medicinal legislation in 2017 but the law did not allow the state until July 2019 to issue patient and caregiver ID cards. Also, the state does not recognize other state medical license cards. *](https://en.wikipedia.org/wiki/Cannabis_in_West_Virginia) |
| [Wyoming](https://en.wikipedia.org/wiki/Wyoming) | ††† | In 2021, bill to legalize cannabis both for recreational and medical use died in the  House of Representatives after it missed a deadline. |

^†^	Powell et al., 2018 (Powell, Pacula, & Jacobson, 2018)

^††^	Martins et al., 2021 (Martins et al.,2021)

^†††^	Cannabis in [State name]. https://en.wikipedia.org/wiki/Cannabis_in_[state name], 2023)

*	Considering either Arkansas as a legalizing jurisdiction as of the end of 2019 or for Ohio, Pennsylvania, North Dakota and West Virginia the year of legalization instead of year of implementation of legalization did not significantly alter the results.
